# Real-time analysis of the cancer genome and fragmentome from plasma and urine short and long cell-free DNA using Nanopore sequencing

**DOI:** 10.1101/2022.08.11.22278674

**Authors:** Ymke van der Pol, Normastuti Adhini Tantyo, Nils Evander, Anouk E. Hentschel, Jip Ramaker, Idris Bahce, Jakko A. Nieuwenhuijzen, Renske D.M. Steenbergen, D. Michiel Pegtel, Norbert Moldovan, Florent Mouliere

## Abstract

Cell-free DNA (cfDNA) can be isolated from blood and/or urine of cancer patients and analyzed with sequencing. Unfortunately, most conventional short-read sequencing methods are technically challenging, labor intensive and time consuming, requiring several days but more typically weeks to obtain interpretable data which are limited by a bias for short cfDNA fragments.

Here, we demonstrate that with Oxford Nanopore Technologies sequencing we can achieve economical and ultra-fast delivery of clinical data from liquid biopsies.

Our ‘ITSFASTR’ approach is able to deliver copy number aberrations, and cfDNA fragmentation profiles in less than 24 hours from sample collection. The tumor-derived cfDNA fraction calculated from lung cancer patient plasma and urine from bladder cancer patients was highly correlated (R=0.98) to the tumor fraction calculated from conventional short-read sequencing of the same samples. cfDNA size profile and fragmentation patterns in plasma and urine exhibited the typical cfDNA features yet with a significantly higher proportion of fragments that exceed 300bp, exhibiting similar tumor fraction than shorter size fragments. Notably, comprehensive fragment-end composition and nucleosome profiling near transcription start sites can be determined from the same data.

We propose that ITSFASTR is the first point-of-care solution for obtaining genomic and fragmentomic results from liquid biopsies.

## Introduction

Cell-free DNA (cfDNA) is intensively investigated as a liquid biopsy in oncology (Heitzer et al., 2019). The genome and epigenome of cancer cells can be non-invasively studied by recovering genetic or epigenetic alterations exhibited by cfDNA fragments (van der Pol and Mouliere, 2019; Wan et al., 2017). Current strategies based on either tumor-naïve or tumor-guided sequencing are reaching high level of sensitivity for detecting tumor-derived signal in plasma of cancer patients (Chabon et al., 2020; Shen et al., 2018; Wan et al., 2020; Zviran et al., 2020). They can also determine and leverage the biological properties of cfDNA (Hudecova et al., 2022; Lo et al., 2021; Mouliere et al., 2018).

Despite their successes, these approaches have limitations for a broad range of applications. They are based on short-read sequencing technologies, inducing a bias towards shorter populations of cfDNA in the bloodstream. They require a complex and expensive sequencing platform, often shared by multiple research groups in genomic facilities, resulting in long waiting time before receiving cfDNA signal data. For a range of applications, for example real-time monitoring of patients under treatment or at home monitoring, more flexible and fast liquid biopsy methods are needed.

Nanopore sequencing using the ONT platform is a highly portable and deployable technology that is capable of sequencing quickly any length of amplified or native DNA fragment or even directly RNA molecules (Euskirchen et al., 2017; Katsman et al., 2022). Due to the higher rate of sequencing error in comparison to short-read sequencers, the ONT platform has rarely been used for cfDNA mutation analysis (Cheng et al., 2015; Marcozzi et al., 2021). However, preliminary works show that broader genomic events, like copy number aberrations, can be recovered and analyzed with ONT Nanopore sequencing of plasma cfDNA (Martignano et al., 2021).

Here we introduce ITSFASTR (InTegrated Sequence and Fragmentome AnalysiS Time Reduction), an ONT sequencing approach that can recover cfDNA somatic copy number aberrations (SCNA) and biological properties with comparable sensitivity to short-read technologies, but with a faster turn-around time. Using ITSFASTR, we generated interpretable genomic copy number aberration, nucleosome profile and fragmentomic data in less than 24 hours from blood collection. We applied the same framework, time frame and sequencing protocol to the analysis of urine cfDNA samples. In addition to the genetic alterations, the ONT platform approach is recovering the long cfDNA fragments, which was previously impossible using short-read technologies, opening a new window into cfDNA biology and structure understanding.

## Results

### Copy number aberrations can be retrieved from the ONT platform data of plasma and urine samples within 24 hours from sampling

To evaluate the potential of ITSFASTR to accurately recover SCNAs from cfDNA, we selected plasma from 12 patients with lung cancer, urine samples from 8 patients with bladder cancer and 2 non-cancer controls (**Figure 1A** and **Table S1**). An aliquot from the same samples was sequenced with a short-read approach for comparison (see Methods). The ONT platform yielded a total of 19,033,791 passed mapping reads, while NovaSeq sequencing a total of 886,395,541 mapping reads (**Table S1**). After sequencing and read processing, SCNA plots were generated with the ichorCNA software (Adalsteinsson et al., 2017). Similar small and large genomic events could be observed in both short and long-read data as illustrated for one patient with lung cancer (**Figure 1A**), and for the whole dataset (**Suppl. Fig. 1**). The amplitude of copy number aberrations per genomic bins was correlated between nanopore and Illumina data, as illustrated in **Figure 1B** for one patient (**Suppl. Fig. 1**). The cfDNA tumor fraction, determined using ichorCNA, was highly correlated between the nanopore and Illumina data (Pearson R=0.98, p<0.001), irrespective of the type of bio-fluid used (plasma or urine) (**Figure 1C**).

**Figure 1:**
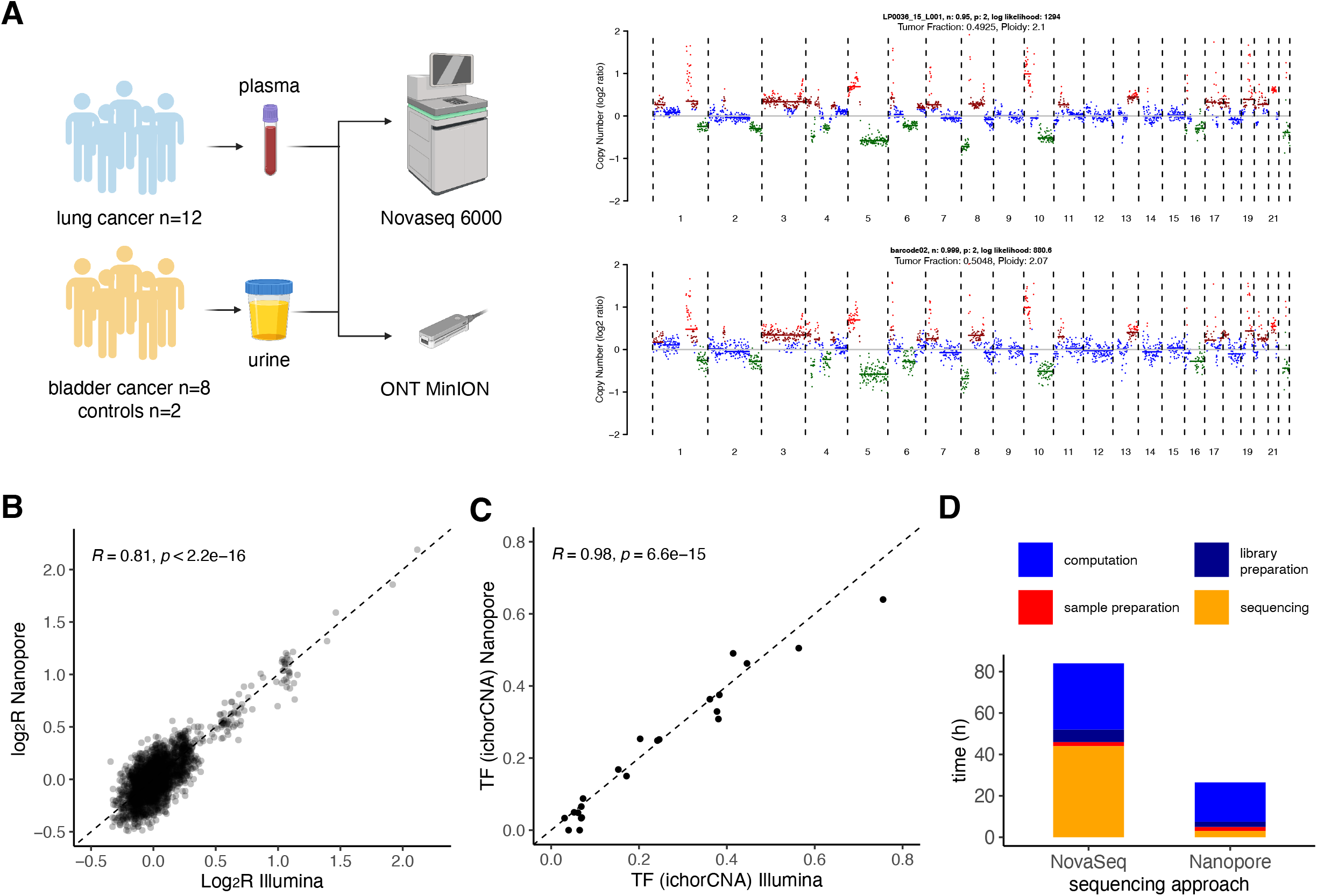
analysis of genetic alterations from plasma and urine cfDNA with the ONT platform. **A:** schematic of the study workflow. 12 plasma samples from lung cancer patients and 10 urine samples from bladder cancer patients (n=8) and controls (n=2) were processed according to an Illumina and Nanopore protocol. Illumina reads were randomly down-sampled to match the read count of the Nanopore reads. Copy number aberration plots were determined using ichorCNA, and exemplified for one patient. **B:** Pearson correlation of the log_2_ratio calculated using ichorCNA per 1M bp genomic bins between a matched sample processed by Illumina sequencing and Nanopore sequencing. **C:** Pearson correlation of the TF calculated using ichorCNA between matched samples processed by Illumina sequencing and Nanopore sequencing. **D:** turn-around time comparison in hours (h) between the NovaSeq and Nanopore protocols from sample collection to generation of interpretable data. The sample preparation phase includes collection, isolation and DNA extraction. The computation phase includes base calling, demultiplexing and data analysis.

The flexibility of the ONT platform allowed fast-turnaround time using an integrated framework like ITSFASTR for liquid biopsy analysis. The time needed from blood collection to interpretable genomic data was ∼24 hours in total (including ∼6 hours of hands-on experiments and ∼18 hours of sequencing and computation time on a laptop with average specifications (see Methods), the main bottle-neck being the base calling step (**Figure 1D**). This can be dramatically reduced to real-time speeds by using a PC with advanced graphical processing unit (GPU) for base calling (Wick et al., 2019), effectively reducing the global turn-around time to ∼7 hours. We calculated the theoretical time needed for generating data on the same samples with a short-read NovaSeq pipeline to ∼84 hours, assuming immediate availability of all required machines and staff to operate them. In practice, the sequencing of an aliquot from the same samples using NovaSeq has taken an average of 34 days (range 31-38 days) from blood collection to interpretable sequencing data due to multiple waiting steps (e.g. before DNA sequencing, for data demultiplexing). The use of ITSFASTR reduced both the theoretical and practical turn-around time to analyze liquid biopsy samples.

### Long cfDNA can be retrieved in the plasma and urine of cancer patients with nanopore sequencing and contains tumor signal

Canonically, cfDNA in blood plasma has been described as short, fragmented molecules centered around 167 bp (and multiple) due to enzymatic cleavage linked to cell-death (Han et al., 2020; van der Pol and Mouliere, 2019). In urine, cfDNA was described as even shorter with a median around 82 bp and the lack of a nucleosome-bound pattern (Markus et al., 2021; Mouliere et al., 2021). This structural organization, observed using short-read sequencing technologies, has been recently challenged by using alternative single-stranded DNA sequencing library preparation (Hudecova et al., 2022), or sequencing technique (Yu et al., 2021).

Using ITSFASTR on plasma from lung cancer patients, the typical nucleosome-bound cfDNA pattern can be observed with a mode of distribution centered around 167 bp and multiple thereof (**Figure 2A** and **Suppl. Fig. 2**). The recovery of long cfDNA (> 300 bp, supposedly linked to di- and tri-nucleosomes) in the plasma of lung cancer patients was increased, as 54.1% of the fragments were longer than 300 bp with the ONT platform compared to 5.3% with short-read sequencing. (**Figure 2A**). The diversity of the trinucleotides at the end of these long cfDNA fragments, determined using Gini index, was not different to the one in shorter (< 300bp) size ranges (**Suppl. Fig. 3**), suggesting that the sequencing was not affecting the entropy of the recovered fragment end sequences. Moreover, long cfDNA fragments (> 300 bp) contained ctDNA tumor signal with an equivalent fraction to shorter cfDNA sizes, as demonstrated following in silico size selection and copy number aberration analysis of long cfDNA (**Figure 2B**). However, we observed a decreased fragment-end diversity for shorter fragments (< 100 bp) (**Suppl. Fig. 3**). To by-pass this limitation, we tested a modification of the ITSFASTR protocol (see Methods) which improved the recovery of high diversity cfDNA fragment-ends in the short size range (**Suppl. Fig. 3**), and mimic the size profile proportions observed using short-read technologies. However, the cfDNA fragment-end sequence composition was altered compared to short-read sequencing. For ITSFASTR the proportion of fragments starting with a T was the highest with a mean of 0.34 compared to 0.32 in the paired short-read dataset, followed by C with a mean of 0.26, compared to 0.27 (**Suppl. Fig. 4**). The ONT protocol used also affected the fragment end sequence composition (**Suppl. Fig. 4**). Beyond cfDNA fragmentation, we evaluated the potential of ITSFASTR to recover other epigenetic features, in particular the nucleosome profiling. We calculated the coverage around a set of known transcription start sites (TSS, n = 6,280 sites) and nucleosome covered quiescent sites, or nucleosome “rich” regions (Quies, n = 8,001) as negative controls using a modified version of Griffin (Doebley et al., 2021). The central coverage in the ± 500 bp region around TSSs was decreased, but as expected not around the Quies regions (**Figure 2D** and **Suppl. Fig. 5-6**). This profile was consistent for the in silico down-sampled short-read and long-read plasma data sets.

**Figure 2.**
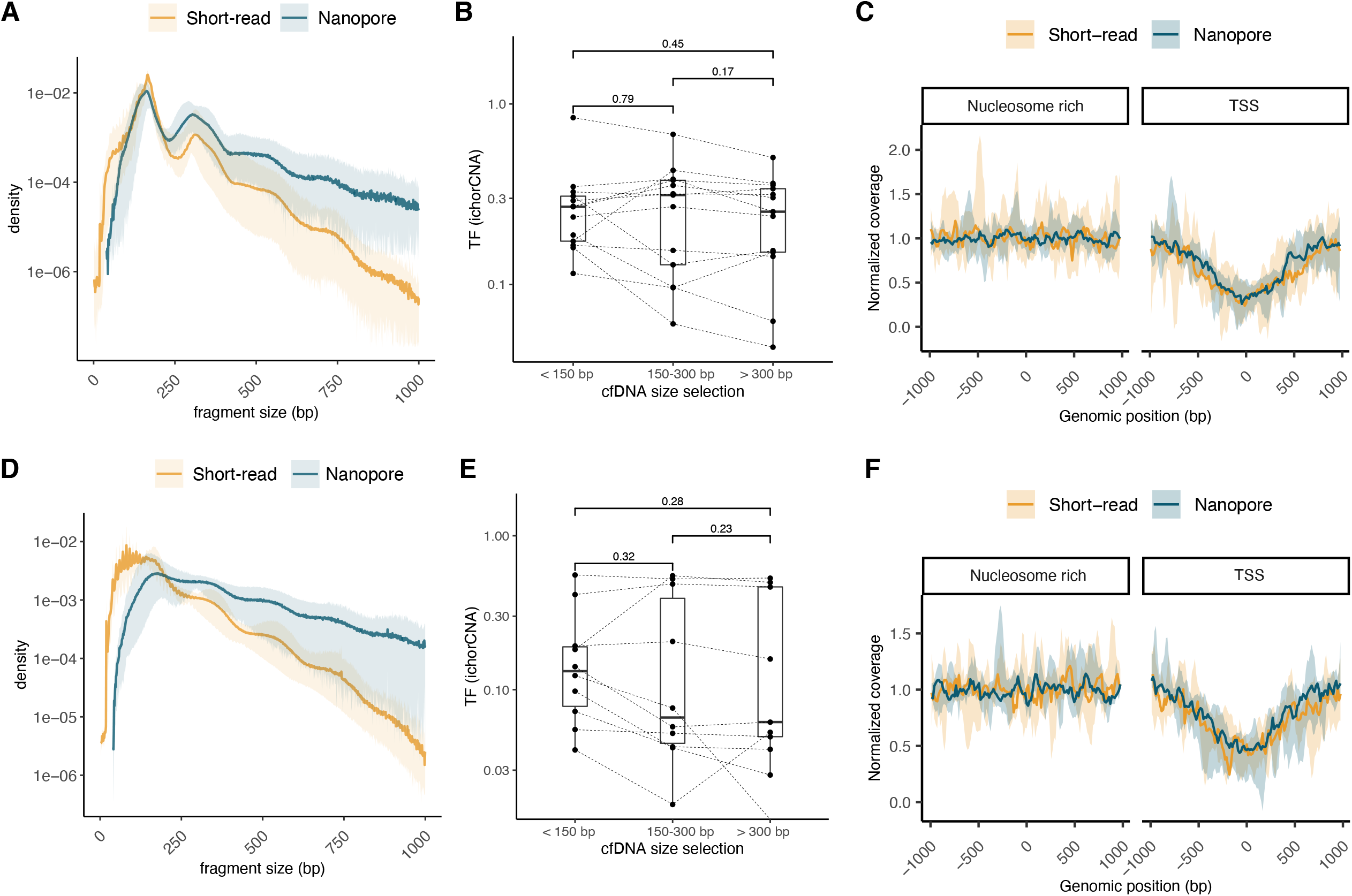
Plasma and urine cfDNA fragmentation patterns using Nanopore and short-read sequencing. In blue are shown Nanopore data and in orange Illumina data. **A:** median density of cfDNA fragment size (bp) from plasma samples with lung cancer (n=14). **B:** tumor fraction (TF) of the plasma samples calculated with ichorCNA depending on specific cfDNA fragment size ranges. P-values calculated using two-sided Wilcoxon test. **C:** normalized coverage of cfDNA from plasma samples of lung cancer patients near TSS regions (±1kbp), and near nucleosome rich regions (±1kbp). Lines showed the median coverage, ribbon the minima and maxima values. **D:** median density of cfDNA fragment size (bp) from urine samples with bladder cancer (n=8). **E:** tumor fraction (TF) of the urine samples calculated with ichorCNA depending on specific cfDNA fragment size ranges. P-values calculated using two-sided Wilcoxon test. **F:** normalized coverage of cfDNA from urine samples of bladder cancer patients near TSS regions (±1kbp), and near nucleosome rich regions (±1kbp). Lines showed the median coverage, ribbon the minima and maxima values.

Applied to urine samples, ITSFASTR recovered cfDNA fragments exhibiting the typical non-nucleosomal patterns previously observed (Markus et al., 2021) using short-read methods for this bio-fluid (**Figure 2D and Suppl. Fig. 7**). However, the median proportion of long cfDNA fragments (> 300 bp) was higher using ITSFASTR in comparison to short-read data (62.4% and 14.2% respectively). Fragments up to 4.28 kbp were detected in the urine from a patient. Similarly, to plasma, > 300 bp cfDNA contained the same fraction of ctDNA tumor signal compared to shorter cfDNA, previously missed by short-read ligation-based sequencing (**Figure 2E** and **Suppl. Fig. 3**). The coverage profiles near TSS and Quies were also consistent between long-read and short-read data from the urine samples (**Figure 2F** and **Suppl. Fig. 8-9**). This demonstrates that ITSFASTR can preserve the nucleosome profile information from liquid biopsy samples.

## Discussion

We introduced an integrated framework (ITSFASTR) based on an ONT sequencing platform that allows analysis of the cancer genome and fragmentome using plasma and urine cfDNA from liquid biopsy in less than 24 hours from sample collection. ITSFASTR generates copy number aberration plots and an estimation of the ctDNA tumor fraction that is highly correlated to what could be achieved using a conventional short-read sequencing framework for liquid biopsy analysis. The capacity of ONT native sequencing to extract a broad range of genomic, epigenomic and transcriptomic features from the same sequencing run is an important advantage to more specialized methods (Euskirchen et al., 2017). ITSFASTR is retaining this feature, by simultaneously generating genomic and fragmentomic data.

Beyond being deployable and having a low starting investment cost, one of the advantages of ITSFASTR is the short turn-around time to deliver interpretable genomic results. In practice, on real-world samples, we were able to recover copy number aberrations and fragmentomic data from blood or urine samples in less than 24 hours, including ∼18 hours of computation. This overall turn-around time could be drastically reduced to ∼7 hours when base calling using a computer with higher-end GPU, producing results in the same day (Wick et al., 2019). Nevertheless, this is an important improvement in comparison to other platforms which in real-world practice are slow to deliver interpretable results.

Beyond this gain in time to deliver results, the ONT sequencing platform (and ITSFASTR by extension) can explore long cfDNA molecules which was previously inefficient using short-read sequencing technologies. A preliminary report using an alternative long-read technology demonstrated the presence of long cfDNA molecules in plasma (Yu et al., 2021). Here, we identify such long fragments (>300 bp) in the plasma and urine of patients with cancer. The sequencing resolution required to recover more detailed fragmentomic signal (e.g. nucleosome occupancy, transcriptomic features on gene level) from such long fragments could be improved by increasing the sequencing time and thus coverage, at the expense of speed, costs and flexibility. Moreover, we observed that these long fragments contain tumor-derived ctDNA signal, which could be critical to recover in clinical conditions when such tumor signal is scarce (e.g. minimal residual disease detection). Contrary to traditional short-read sequencing approaches, the full length of the molecule is sequenced for every fragment size with the ONT sequencing platform, which can provide useful information about the allelic occurrence of mutations, or phasing (Suzuki et al., 2017). Using short-read techniques, phased variants have shown potential to improve the sensitivity to detect minutes amount of tumor DNA in plasma (Kurtz et al., 2021), but the potential of long-read techniques remains untapped.

Fragmentation-based nucleosome profiling from plasma samples is a promising alternative to more conventional and labor-intensive methods for the deconvolution of chromatin states in the genome (Lo et al., 2021; Snyder et al., 2016). We demonstrated that the nucleosome position profile is preserved and can be retrieved from data produced by ITSFASTR. Such information can be retrieved from various bio-fluids, including plasma and urine samples. Nucleosome profiling enabled tissue-of-origin deconvolution from plasma samples of high depth WGS data (Snyder et al., 2016; Zhu et al., 2021). Thus, ITSFASTR could potentially power ultra-fast nucleosome-profile based tissue-of-origin deconvolution.

The sample size of this study is small and will require further confirmation with larger cohorts of bio-fluids for validation in clinical situations. In particular, an application to clinical situation where high intensity sampling (e.g. multiple samples per week for follow-up post-surgery, or analysis of diurnal modifications) could help to demonstrate the potential of the approach. Per nature, ONT sequencing was used in a low coverage whole genome sequencing (lc-WGS) mode which is affected by the same limitation in terms of sensitivity as lc-WGS from short-read technologies (Adalsteinsson et al., 2017; Heitzer et al., 2013). Although not affecting the detection of large genomic events or fragmentomic analysis, one drawback of ONT sequencing is the single read base accuracy, which is currently inferior to that of conventional short-read approaches, and can affect SNV calling. Alternative approaches are under development to improve the recovery of SNV from plasma cfDNA using ONT sequencing (Marcozzi et al., 2021). Moreover, the pre-analytical conditions that are affecting cfDNA short-read sequencing and fragmentomic analysis could alter differently longer cfDNA fragments (van der Pol et al., 2022).

In conclusion, same-day delivery of genomic and fragmentomic signatures from plasma and urine cfDNA is possible via an integrated nanopore liquid biopsy framework. Using ITSFASTR, a novel framework for liquid biopsy analysis, we identify longer fragments of cfDNA in the plasma but also urine of patients with cancer, which were not previously observed. We determined these long fragments contained tumor-derived ctDNA molecules that could be important to recover for some liquid biopsy applications.

## Material and Methods

### Study design

A total of 12 plasma and 10 urine samples from 22 individuals were retrieved across 2 cancer types (**Table S1**). Lung cancer patients were recruited following informed consent via the Liquid Biopsy Center at the Amsterdam UMC, location VUmc and location AMC (study approved by the Amsterdam UMC ethics board, METC U2019_035). Bladder cancer patients were recruited following informed consent at the Amsterdam UMC, location VUmc (study approved by the Amsterdam UMC ethics board, METC 2018.355).

### Sample preparation

Blood samples were collected before treatment in EDTA K2 coated tubes and processed using a double-centrifugation protocol (900 g for 15 minutes; 2500 g for 10 minutes at room temperature). Supernatant plasma was aliquoted in 0.5mL Nunc tubes before being stored at -80°C. Urine samples were collected before cystoscopy or transurethral resection of the bladder tumor and were processed within 24 to 72 h. DNA quality was preserved by the addition of 0.6 M EDTA to the collection tube in a final concentration of 40 mM. Urine samples were pelleted by centrifugation of 15 mL urine at 800 g for 10 min. The urine supernatant was collected and thereafter stored at -□80 °C. cfDNA from lung cancer plasma and bladder cancer urine samples were extracted using the automated system QiaSymphony Circulating DNA Kit (Qiagen) with 2 to 3.2 mL of sample input (PBS was added when required to reach 3.2 mL). Six matched plasma samples were extracted using MagMAX Cell-Free DNA Isolation Kit (Thermo Fisher) according to the manufacturer’s instructions with 1 mL plasma input. cfDNA concentration was measured using Cell-free DNA ScreenTape Analysis of the Agilent 4200 TapeStation System (Agilent).

### Sequencing preparation

For short-reads sequencing, ThruPLEX Plasma-seq Kit (Takara Bio) was used for library preparation and sequenced after equimolar pooling using 150-bp paired-end mode on NovaSeq 6000 (Illumina) with S4 flow cells. Long-read sequencing was performed using R9.4.1 flow cells on a MinION device (Oxford Nanopore Technologies). Two plasma sample libraries were prepared using the PCR Barcoding Kit (SQK-PBK004, Oxford Nanopore Technologies) according to the manufacturer’s instructions (Protocol 1). This protocol was adapted to short cfDNA (Protocol 2) by increasing the ethanol concentration to 80% on all washing steps and the amount of PCR cycles was set to 18 cycles to account for the low amount of cfDNA input. The beads-to-sample ratio was also altered to 1.8X according to a previously published study (Martignano et al., 2021). A second optimization was done by reverting the beads-to-sample ratio to the original SQK-PBK004 protocol while keeping the amount of PCR cycles at 18 and ethanol concentration at 80% (Protocol 3). The quality of libraries was checked using the D1000 ScreenTape Analysis Assay of the Agilent 4200 TapeStation System (Agilent) prior to pooling. Libraries were pooled in equimolar amounts with 50-100 fmoles as the total input for sequencing.

### Base calling and demultiplexing

Base calling of the short-read sequencing was performed using the NovaSeq control software (v. 1.7.5). Demultiplexing was performed running bcl2fastq2 (v. 2.20). Nanopore reads were base called and demultiplexed on a laptop with Intel i7-7500U/BGA quad core processor, 8 GB DDR4 RAM, running Ubuntu LTS 20.04 and MinKNOW (v. 22.03.6), with Guppy (v. 6.0.7) running in CPU Fast mode.

### Read preprocessing

Short reads were trimmed using bbduk (bbmap v. 38.79 commit h516909a_0) with the ktrim=r, k=23, mink=11, hdist=1 parameters and default adapter sequences provided with the tool. Mapping to the human reference genome (GRCh38) was performed using BWA MEM (v. 0.7.17 commit hed695b0_7) with default parameters. Long reads were trimmed with Porechop (v. 0.2.4) with default parameters and adaptor sets, except extra_end_trimming, which was set to 0. Trimmed long reads were mapped to the human reference genome (GRCh38) using Minimap2 (v. 2.24) with the ax map-ont parameter. Duplicate reads from both short- and long-alignments were marked using Sambamba (v. 0.8.1) with default parameters. Unmapped reads, secondary or supplementary alignments, PCR duplicates and alignments with a quality < 5 were removed using Samtools (v. 1.12). Short-read datasets were down-sampled to the same coverage as the long-read data using Samtools (v. 1.12) view -bs i.p where i=95, a randomly generated number, while p was the proportion of long-reads compared to the number of short-reads.

### Copy number analysis and tumor fraction estimation

The ichorCNA software (v. 0.3.2.0) was used to perform the copy number analysis and estimate the ctDNA tumor fraction for both short- and long-read data (Adalsteinsson et al., 2017). Exceptions to the software’s default settings are as follows: (1) An in-house panel-of-normals from shallow Whole Genome Sequencing was created; (2) non-tumor fraction parameter restart values were increased to c(0.95,0.99,0.995,0.999); (3) ichorCNA ploidy parameter restart value was set to 2; (4) no states were used for subclonal copy number and (5) the maximum copy number to use was lowered to 3. The tumor fraction with the highest log likelihood was retrieved and reported.

### Fragmentomic analysis

cfDNA fragment length was retrieved from the mapped short reads using Picard (v. 2.22.2) CollectInsertSizeMetrics with default parameters. Fragment lengths for the mapped long reads were computed using NanoPlot (v. 1.40.0) with the *alength, raw* and *huge* parameters. Fragment-end sequence proportions of the aligned reads were retrieved and their Gini index was calculated using our Fragment End Integrated Analysis (FrEIA) tool (commit 8ecb58b) [https://github.com/mouliere-lab/FrEIA], with the *mode* parameter set to either *Illumina* for short reads or *ONT* for long reads, and FragmSizeMax set to 5000 for the long-read data. Fragmentation-based nucleosome positioning analysis was performed using a modified version of Griffin (commit <u>73c605a</u>) [https://github.com/adoebley/Griffin] called Griffin-LRS (Doebley et al., 2021). We modified the original code by adding the possibility of working with single-end nanopore sequencing. We run Griffin-LRS with the original parameters, except setting *map_quality* to 5 for both *griffin_GC_and_mappability_correcction* and *griffin_nucleosome_profiling*. GRCh38 was used as the reference genome. For the target sites we retreaved the TSS positions for known genes (TssA regions) and nucleosome covered regions (quies regions) from the UCSC website (https://hgdownload.soe.ucsc.edu/downloads.html#human) and filtered sites with low mappability, using *griffin_filter_sites*.

## Data Availability

Sequencing data will be deposited at the EGA upon acceptance of the manuscript.

## Data availability

Sequencing data will be deposited at the EGA upon acceptance of the manuscript.

## Author contributions

Conception and design: YP, NM, FM

Experiments and data collection: YP, NAT

Data processing: YP, NM

Software development: NM

Data analysis: YP, NAT, NM, FM

Sample acquisition: RS, JN, IB

Funding acquisition: DMP, FM

Manuscript draft: YP, NAT, NM, FM

Manuscript revisions and comments: YP, NAT, NE, JN, IB, RS, DMP, FM

Supervision: FM

## Acknowledgments

The authors would like to thank Dr. Trang Vu and Dr. Daoud Sie for their help and constructive discussions. The authors are thankful to the Amsterdam UMC Liquid Biopsy Center for the logistical support. Y.P. and F.M. are funded by the Amsterdam UMC Liquid Biopsy Center, an initiative made possible through the Stichting Cancer Center Amsterdam. N.M. and F.M. are supported by a Dutch Cancer Fund (KWF-12822). This work was carried out on the Dutch national e-infrastructure with the support of SURF Cooperative.

