## Supplementary Figures for "Real-time analysis of the cancer genome and fragmentome from plasma and urine short and long cell-free DNA using Nanopore sequencing"

#### Supplementary Figure 1

Patient 1

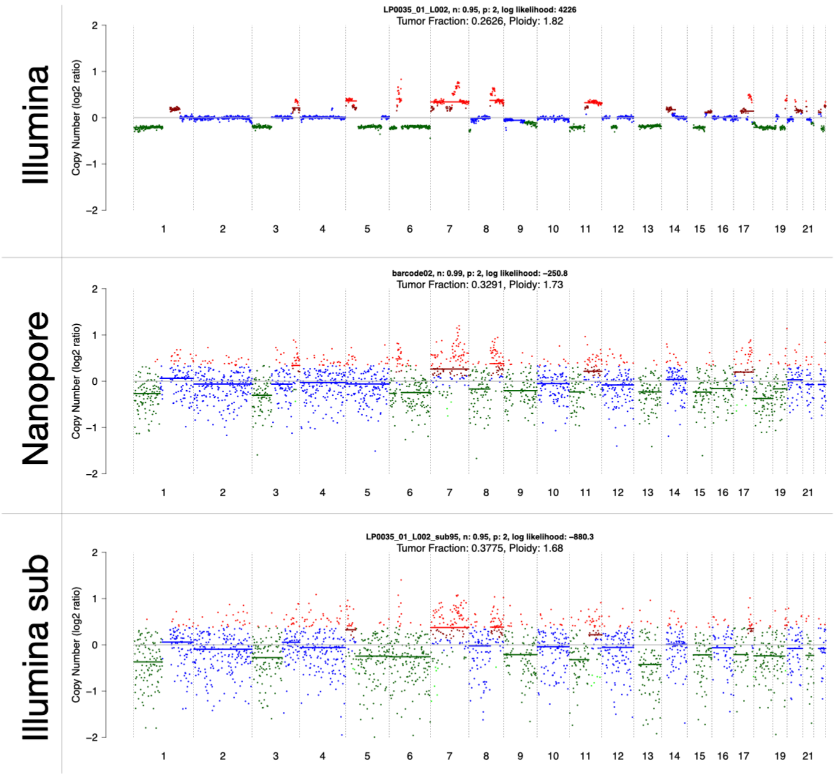


Patient 98

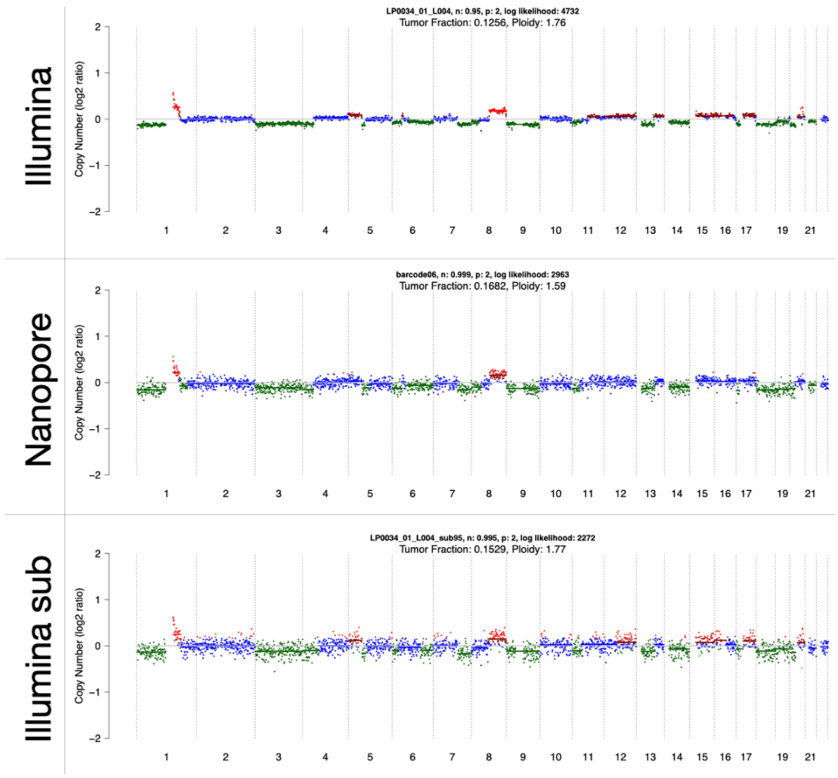
Illumina
LP0034_01_L004, n: 0.95, p: 2, log likelihood: 4732
Tumor Fraction: 0.1256, Ploidy: 1.76
Copy Number (log2 ratio)
2
1
0
−1
−2
1 2 3 4 5 6 7 8 9 10 11 12 13 14 15 16 17 19 21
Nanopore
barcode06, n: 0.999, p: 2, log likelihood: 2963
Tumor Fraction: 0.1682, Ploidy: 1.59
Copy Number (log2 ratio)
2
1
0
−1
−2
1 2 3 4 5 6 7 8 9 10 11 12 13 14 15 16 17 19 21
Illumina sub
LP0034_01_L004_sub95, n: 0.995, p: 2, log likelihood: 2272
Tumor Fraction: 0.1529, Ploidy: 1.77
Copy Number (log2 ratio)
2
1
0
−1
−2
1 2 3 4 5 6 7 8 9 10 11 12 13 14 15 16 17 19 21

Patient 163

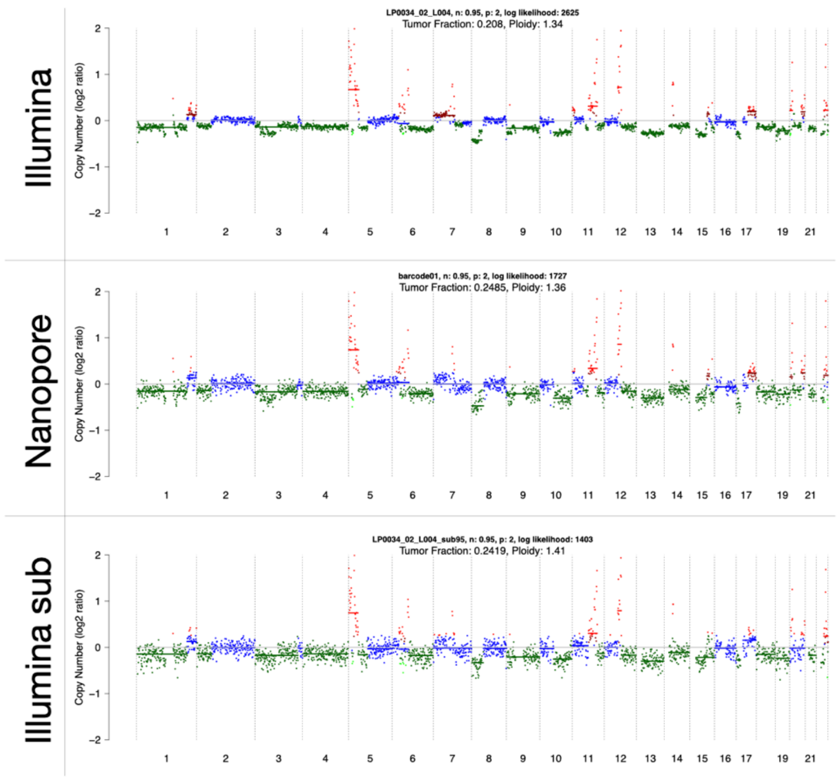
Illumina
LP0034_02_L004, n: 0.95, p: 2, log likelihood: 2625
Tumor Fraction: 0.208, Ploidy: 1.34
Copy Number (log2 ratio)
2
1
0
−1
−2
1 2 3 4 5 6 7 8 9 10 11 12 13 14 15 16 17 19 21
Nanopore
barcode01, n: 0.95, p: 2, log likelihood: 1727
Tumor Fraction: 0.2485, Ploidy: 1.36
Copy Number (log2 ratio)
2
1
0
−1
−2
1 2 3 4 5 6 7 8 9 10 11 12 13 14 15 16 17 19 21
Illumina sub
LP0034_02_L004_sub95, n: 0.95, p: 2, log likelihood: 1403
Tumor Fraction: 0.2419, Ploidy: 1.41
Copy Number (log2 ratio)
2
1
0
−1
−2
1 2 3 4 5 6 7 8 9 10 11 12 13 14 15 16 17 19 21

Patient 215

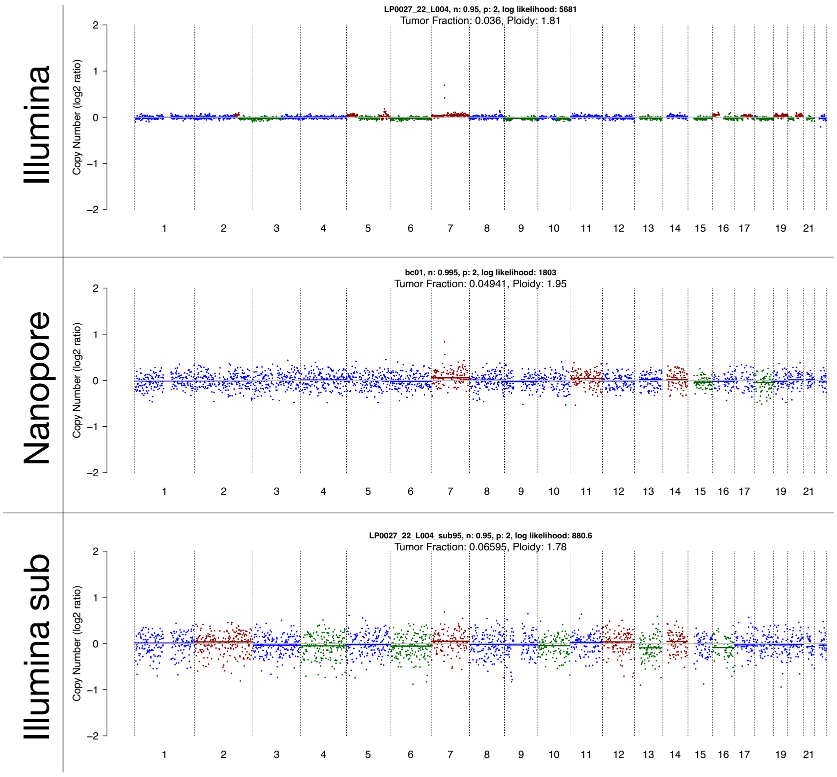


Patient 254

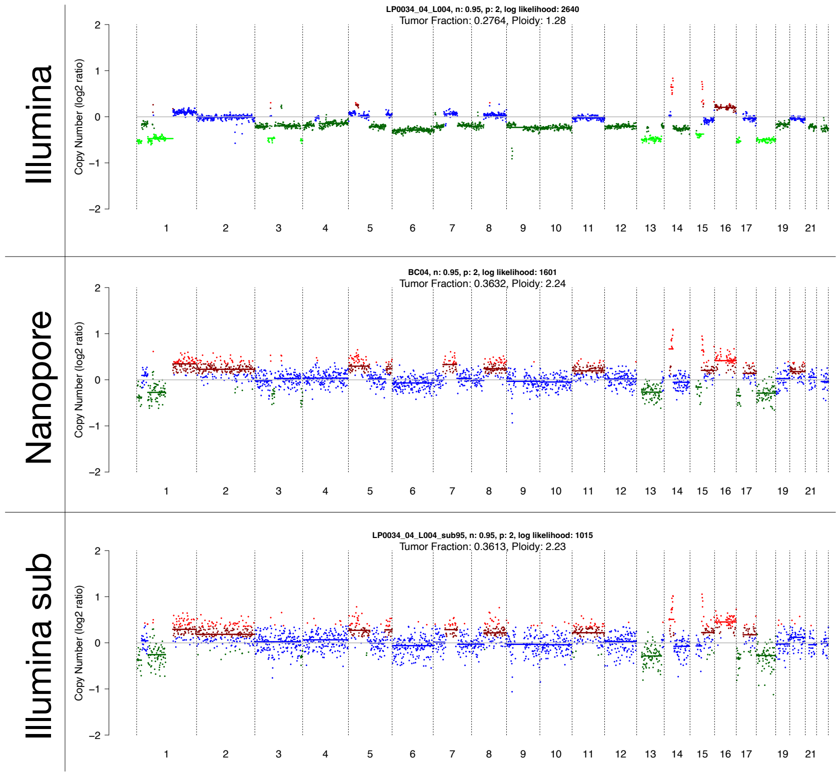
Illumina
LP0034_04_L004, n: 0.95, p: 2, log likelihood: 2640
Tumor Fraction: 0.2764, Ploidy: 1.28
Copy Number (log2 ratio)
Nanopore
BC04, n: 0.95, p: 2, log likelihood: 1601
Tumor Fraction: 0.3632, Ploidy: 2.24
Copy Number (log2 ratio)
Illumina sub
LP0034_04_L004_sub95, n: 0.95, p: 2, log likelihood: 1015
Tumor Fraction: 0.3613, Ploidy: 2.23
Copy Number (log2 ratio)

Patient 298

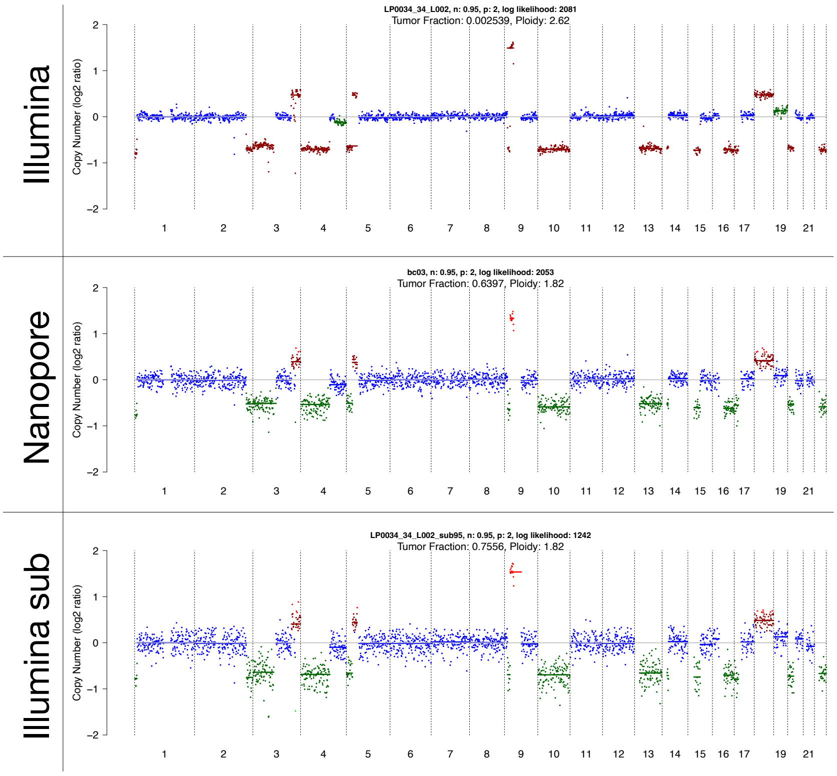
Illumina
LP0034_34_L002, n: 0.95, p: 2, log likelihood: 2081
Tumor Fraction: 0.002539, Ploidy: 2.62
Nanopore
bc03, n: 0.95, p: 2, log likelihood: 2053
Tumor Fraction: 0.6397, Ploidy: 1.82
Illumina sub
LP0034_34_L002_sub95, n: 0.95, p: 2, log likelihood: 1242
Tumor Fraction: 0.7556, Ploidy: 1.82
Copy Number (log2 ratio)

### Patient 321

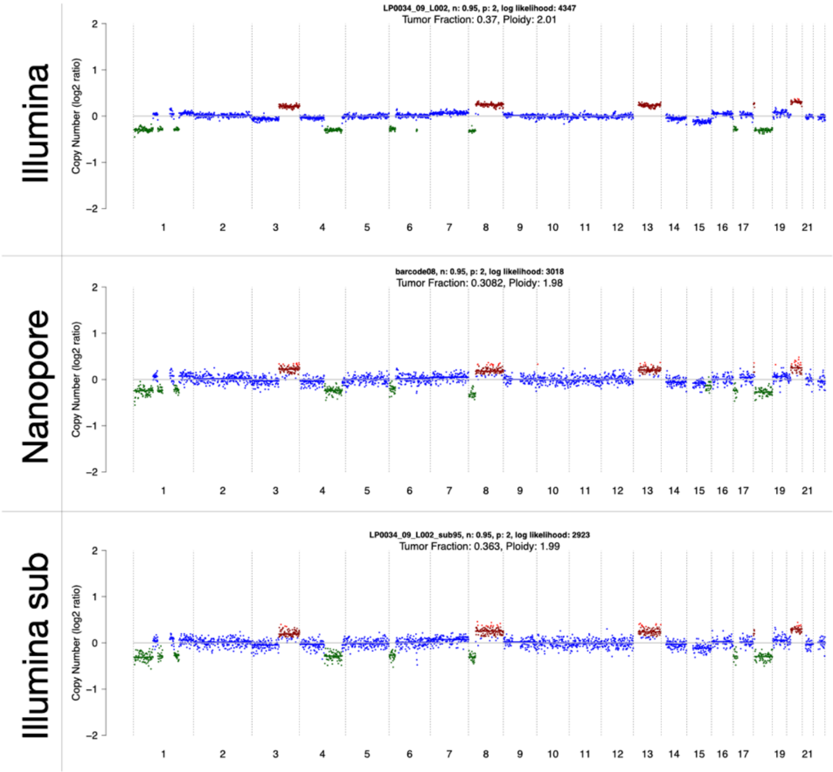
Illumina
LP0034_09_L002, n: 0.95, p: 2, log likelihood: 4347
Tumor Fraction: 0.37, Ploidy: 2.01
Nanopore
barcode08, n: 0.95, p: 2, log likelihood: 3018
Tumor Fraction: 0.3082, Ploidy: 1.98
Illumina sub
LP0034_09_L002_sub95, n: 0.95, p: 2, log likelihood: 2923
Tumor Fraction: 0.363, Ploidy: 1.99
Copy Number (log2 ratio)

#### Patient 522

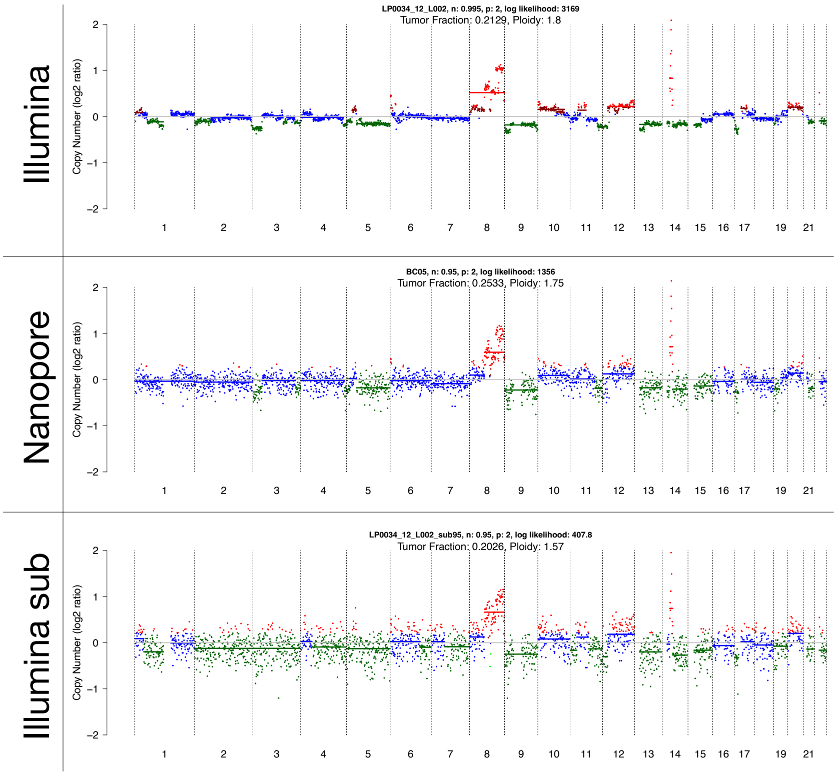


#### Patient 539

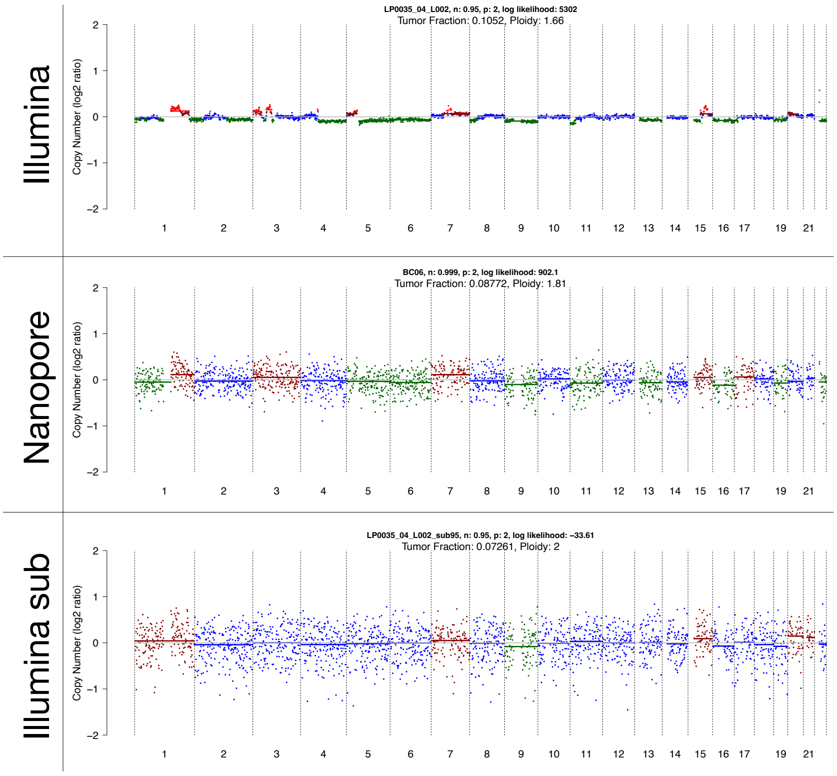
Illumina
LP0035_04_L002, n: 0.95, p: 2, log likelihood: 5302
Tumor Fraction: 0.1052, Ploidy: 1.66
Nanopore
BC06, n: 0.999, p: 2, log likelihood: 902.1
Tumor Fraction: 0.08772, Ploidy: 1.81
Illumina sub
LP0035_04_L002_sub95, n: 0.95, p: 2, log likelihood: −33.61
Tumor Fraction: 0.07261, Ploidy: 2
Copy Number (log2 ratio)
1 2 3 4 5 6 7 8 9 10 11 12 13 14 15 16 17 19 21

Figure 614

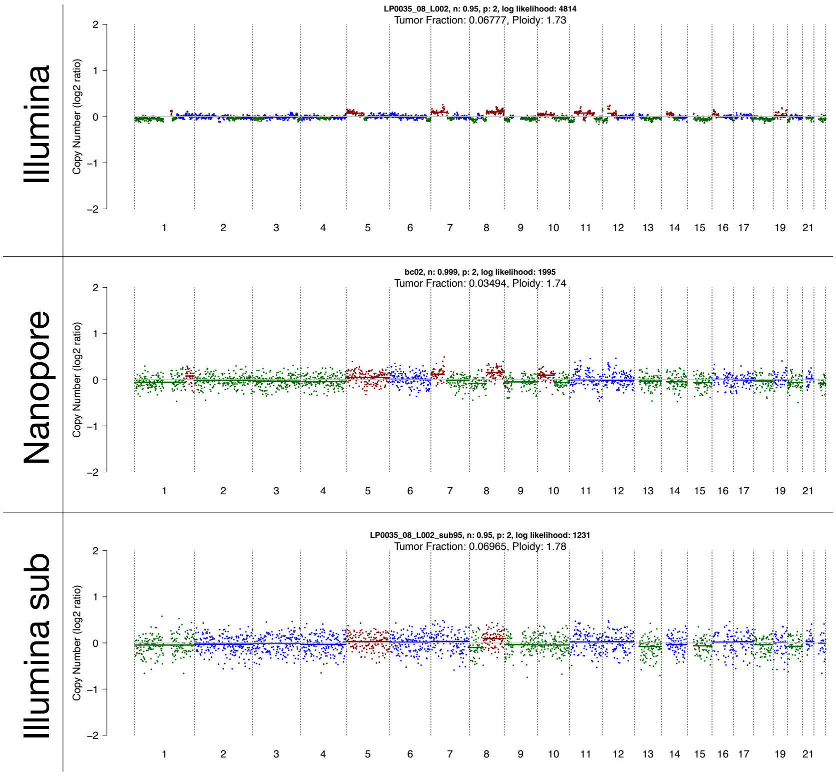
Illumina
LP0035_08_L002, n: 0.95, p: 2, log likelihood: 4814
Tumor Fraction: 0.06777, Ploidy: 1.73
Nanopore
bc02, n: 0.999, p: 2, log likelihood: 1995
Tumor Fraction: 0.03494, Ploidy: 1.74
Illumina sub
LP0035_08_L002_sub95, n: 0.95, p: 2, log likelihood: 1231
Tumor Fraction: 0.06965, Ploidy: 1.78
Copy Number (log2 ratio)
1 2 3 4 5 6 7 8 9 10 11 12 13 14 15 16 17 19 21

#### Patient 685

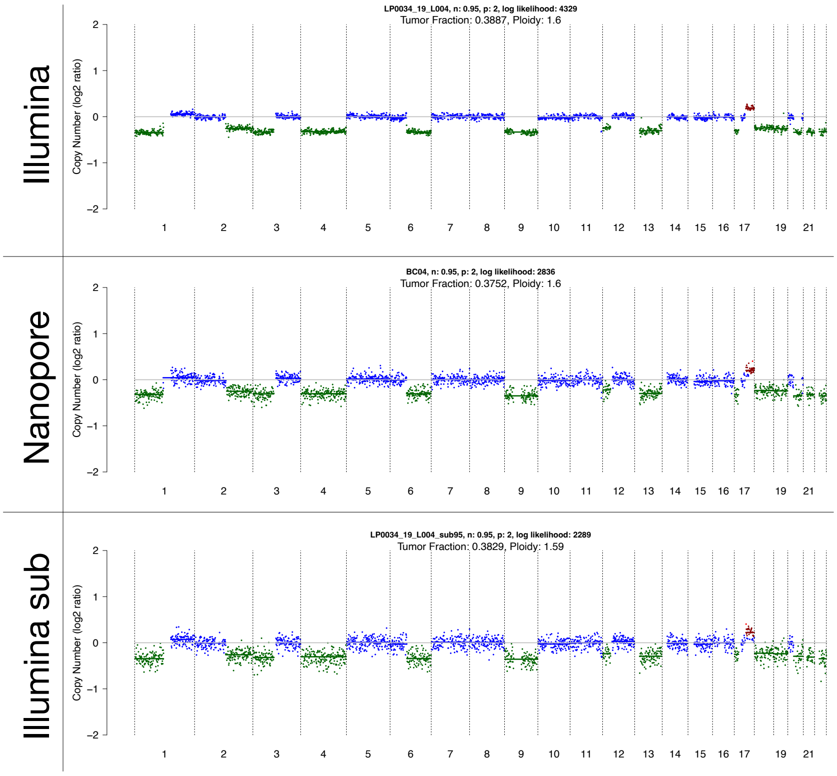


#### Patient 690

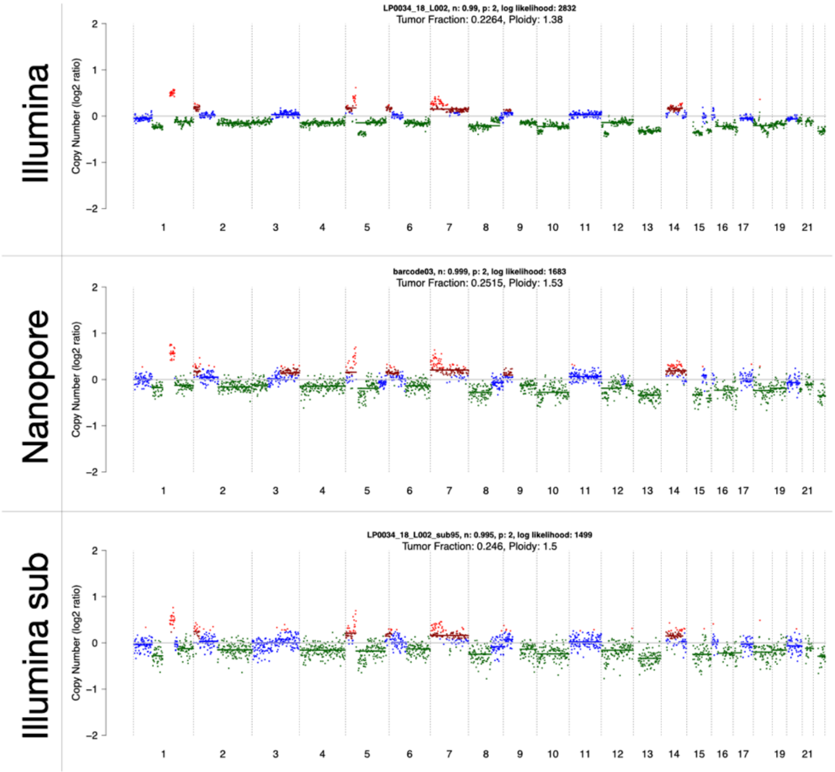

#### Patient A130E

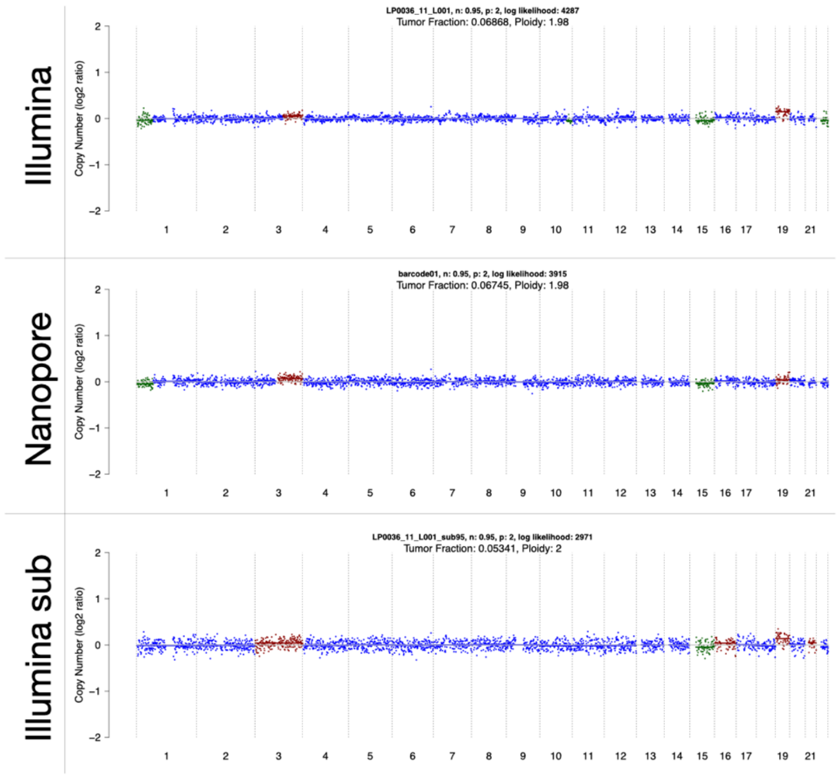


#### Patient A136E

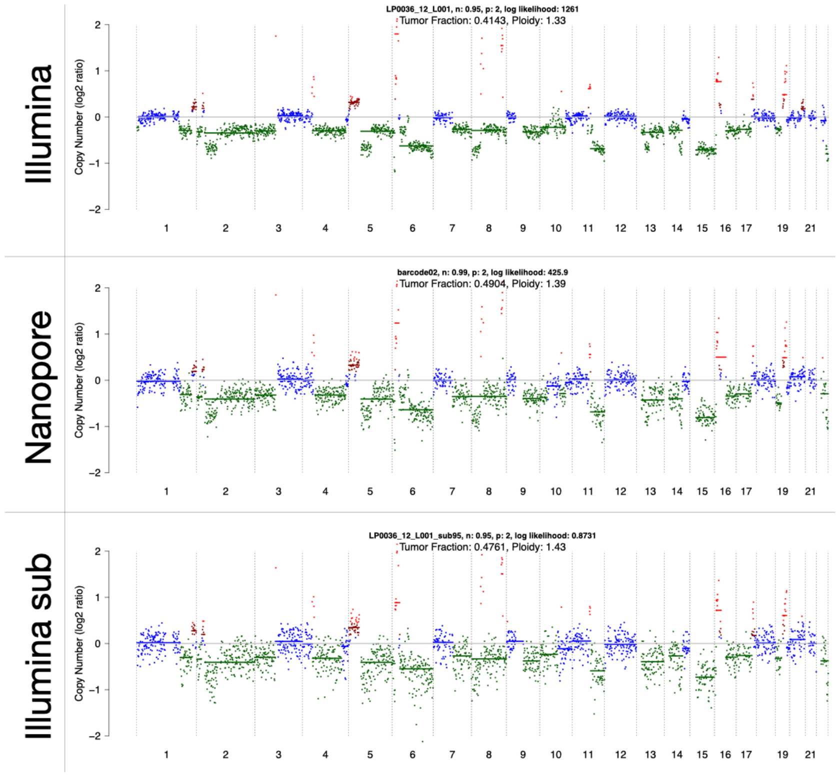

Patient A146E

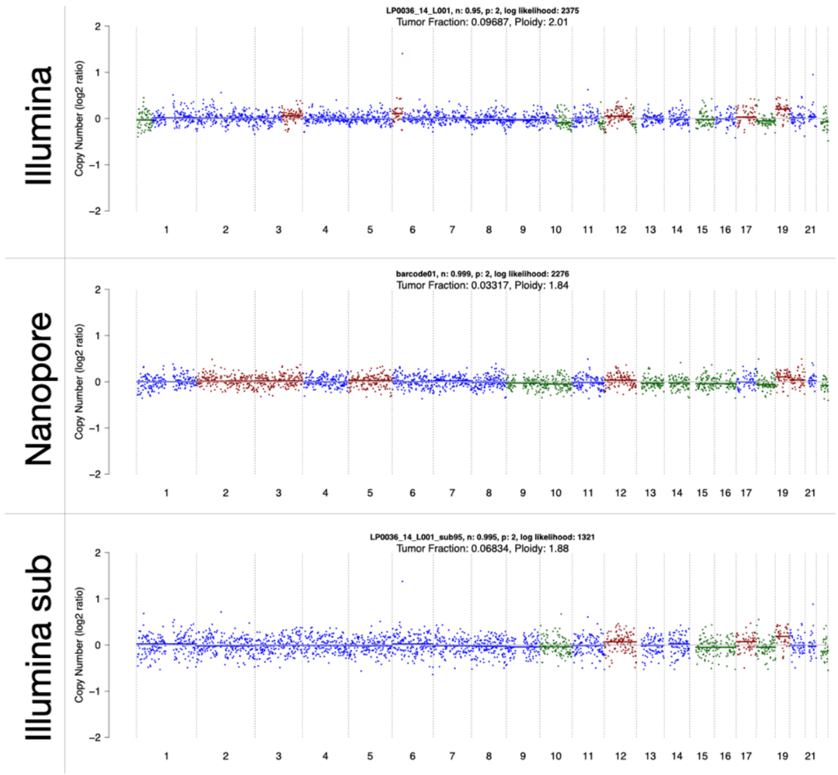


Patient A151E

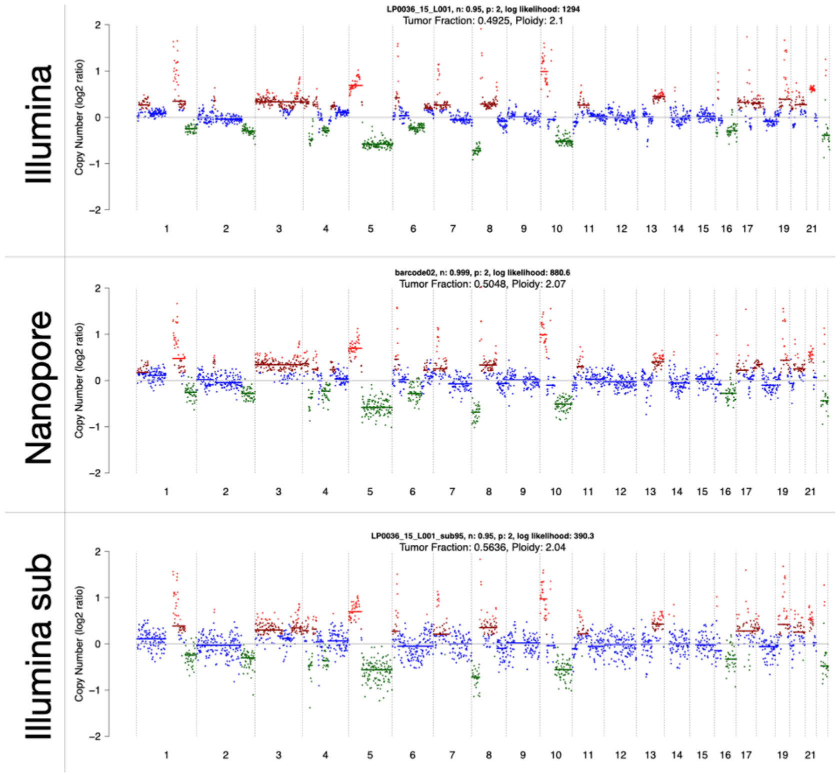

Patient A155E

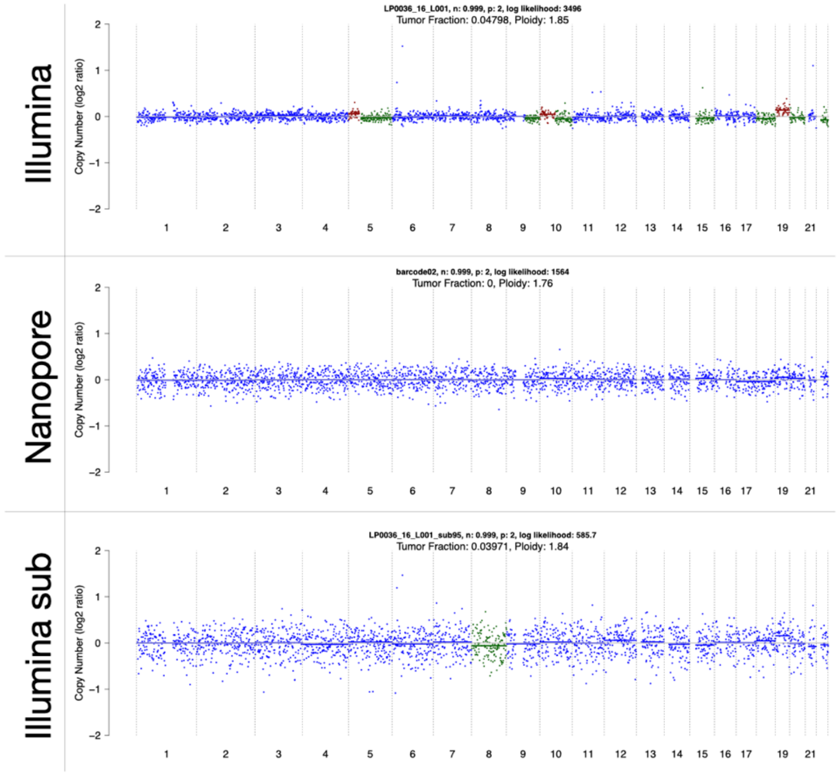


Patient A163E

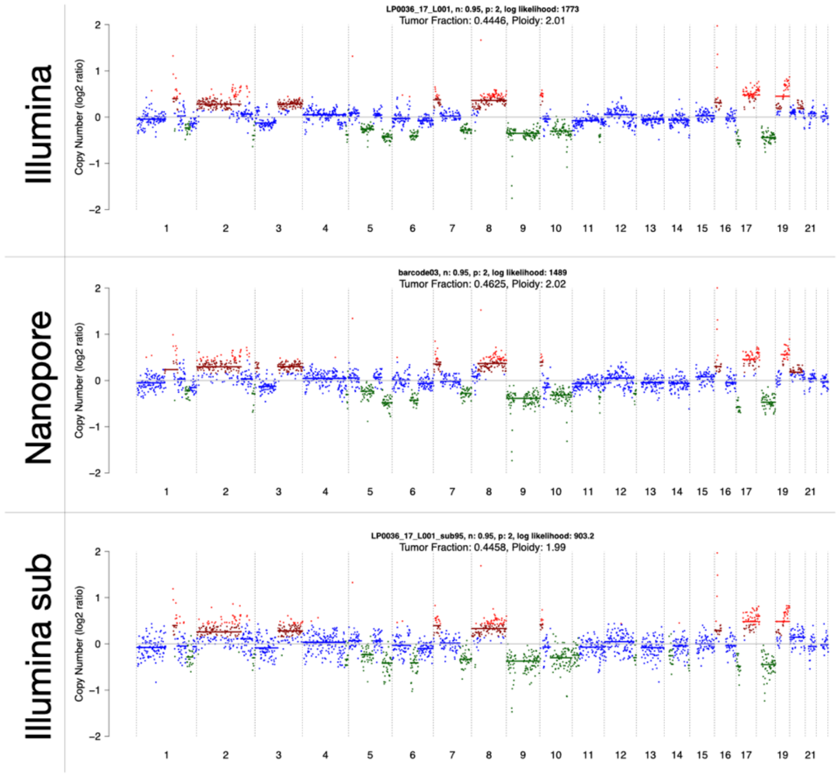

Patient D29

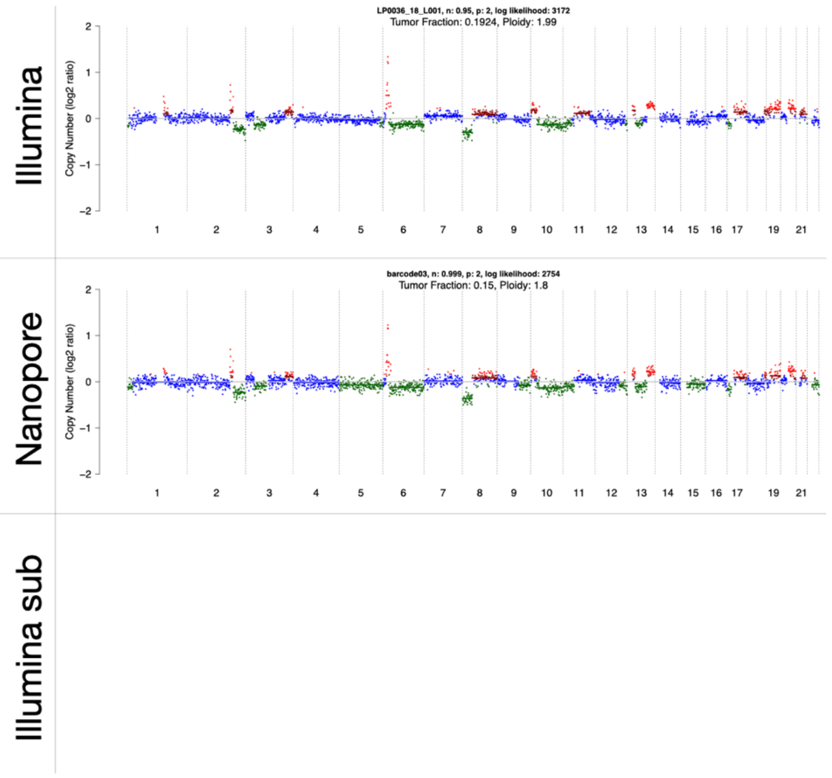


Patient D101

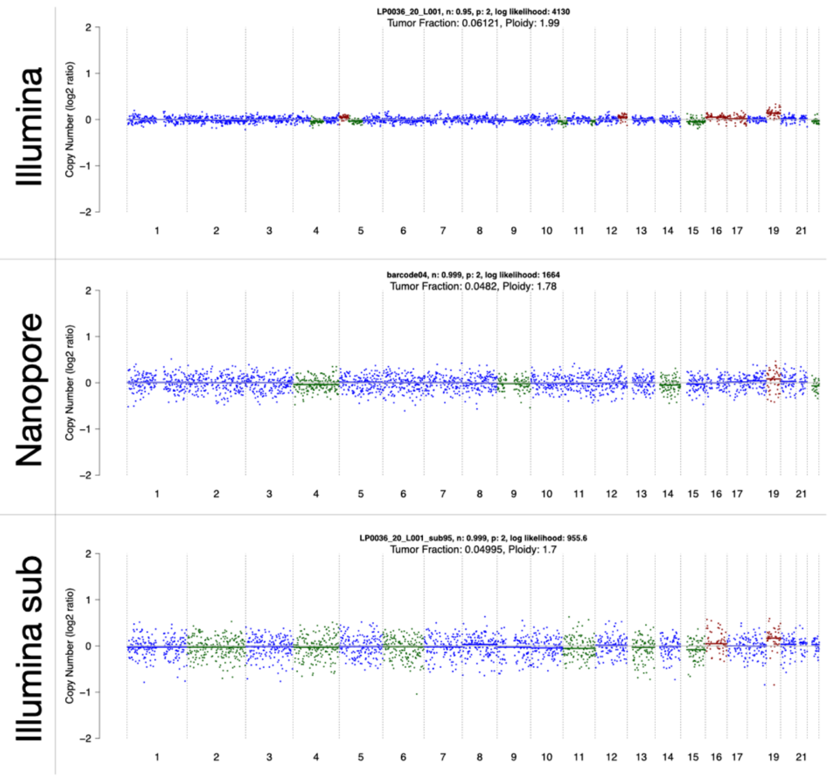

Patient N123

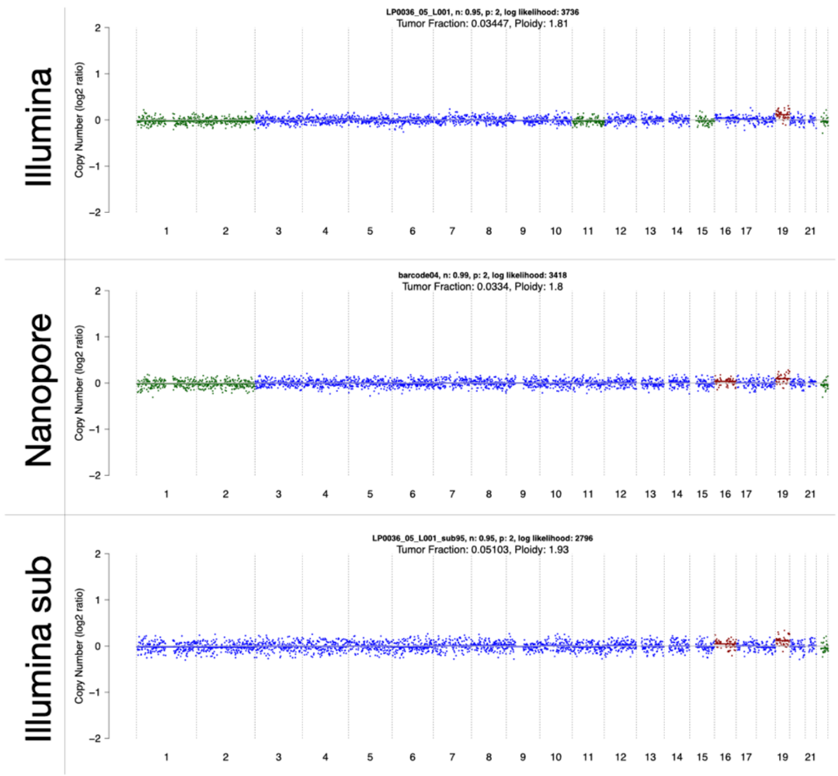


Patient N195E

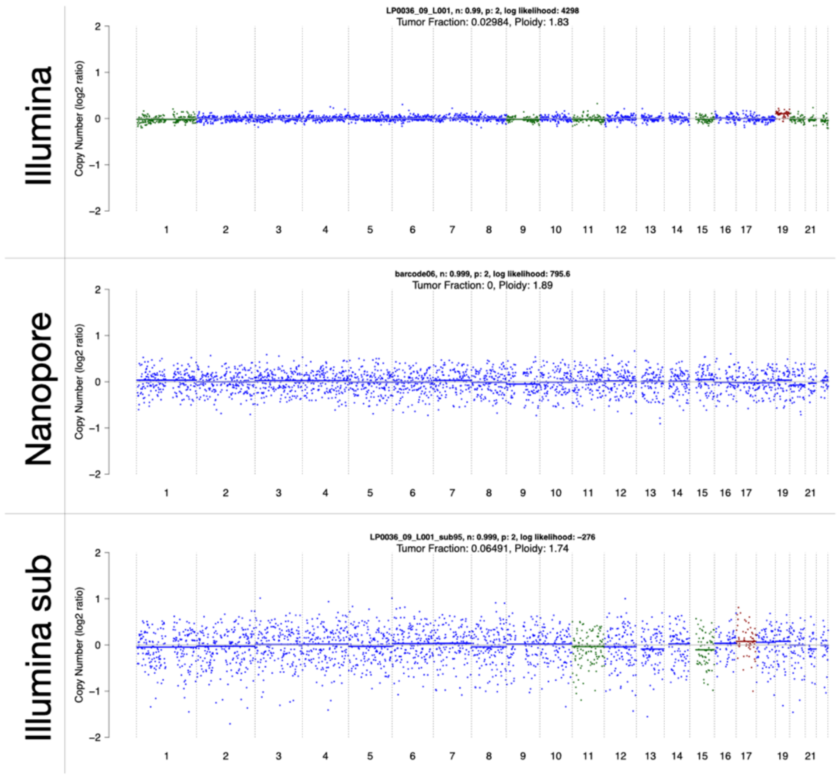

**Supplementary Figure 1. Copy number aberration profiles for the samples included in this study.** For each sample the upper panel correspond to the data from the Illumina short-read sequencing, the middle panel to the ONT sequencing and the bottom panel to the Illumina sequencing down-sampled randomly to the same number of reads as the ONT data.

**Supplementary Figure 2**

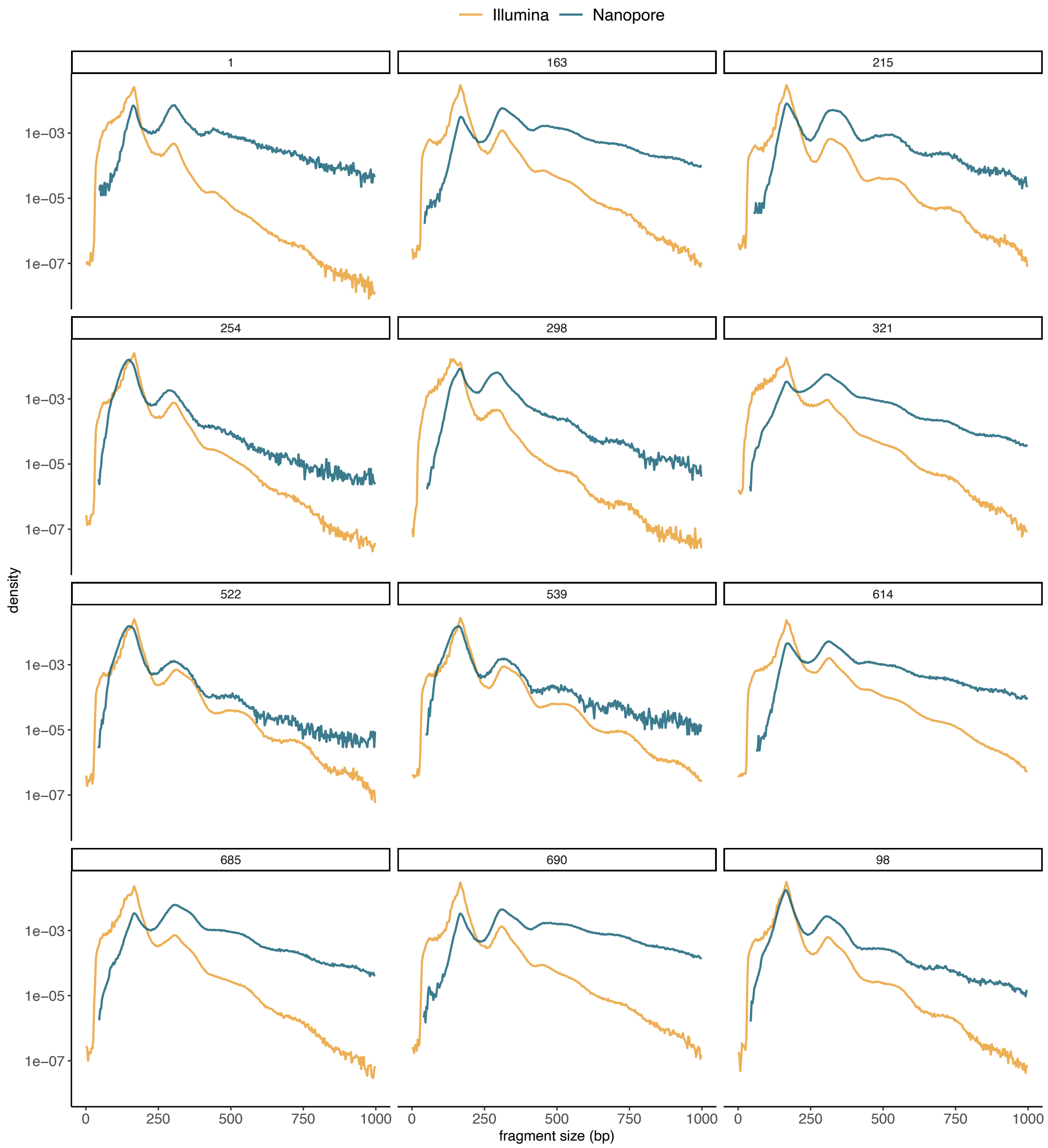


**Supplementary Figure 2. cfDNA fragment size distribution for the plasma samples included in this study.** Color indicates the sequencing type (blue: Nanopore and orange: Illumina). Header numbers represent patient IDs.

**Supplementary Figure 3**

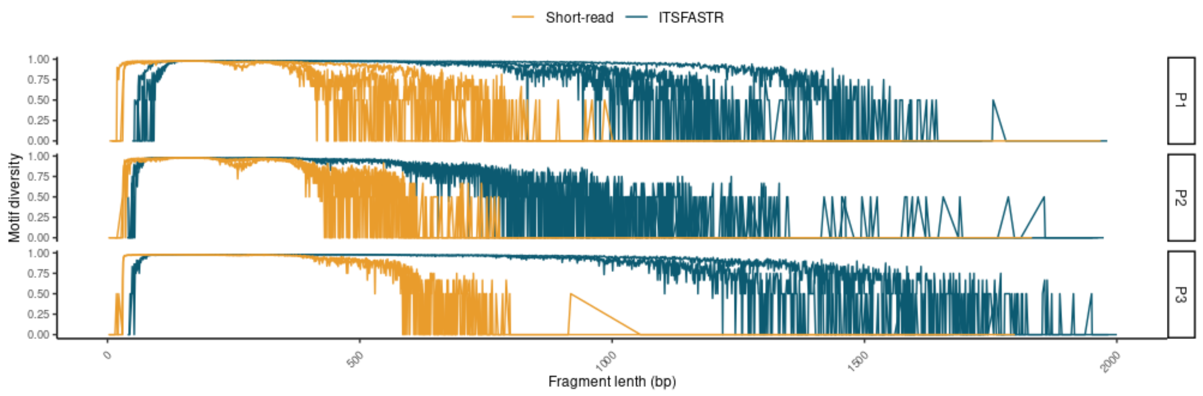


**Supplementary Figure 3. cfDNA fragment-end motif diversity calculated for each protocol tested in the study.** Color indicates the sequencing type (blue: Nanopore and orange: Illumina).

#### Supplementary Figure 4

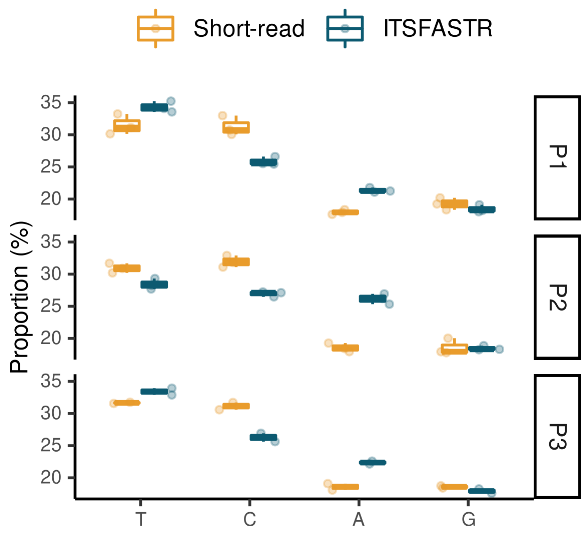


**Supplementary Figure 4. cfDNA fragment-end mononucleotide proportions calculated for each protocol tested in the study.**

#### Supplementary Figure 5

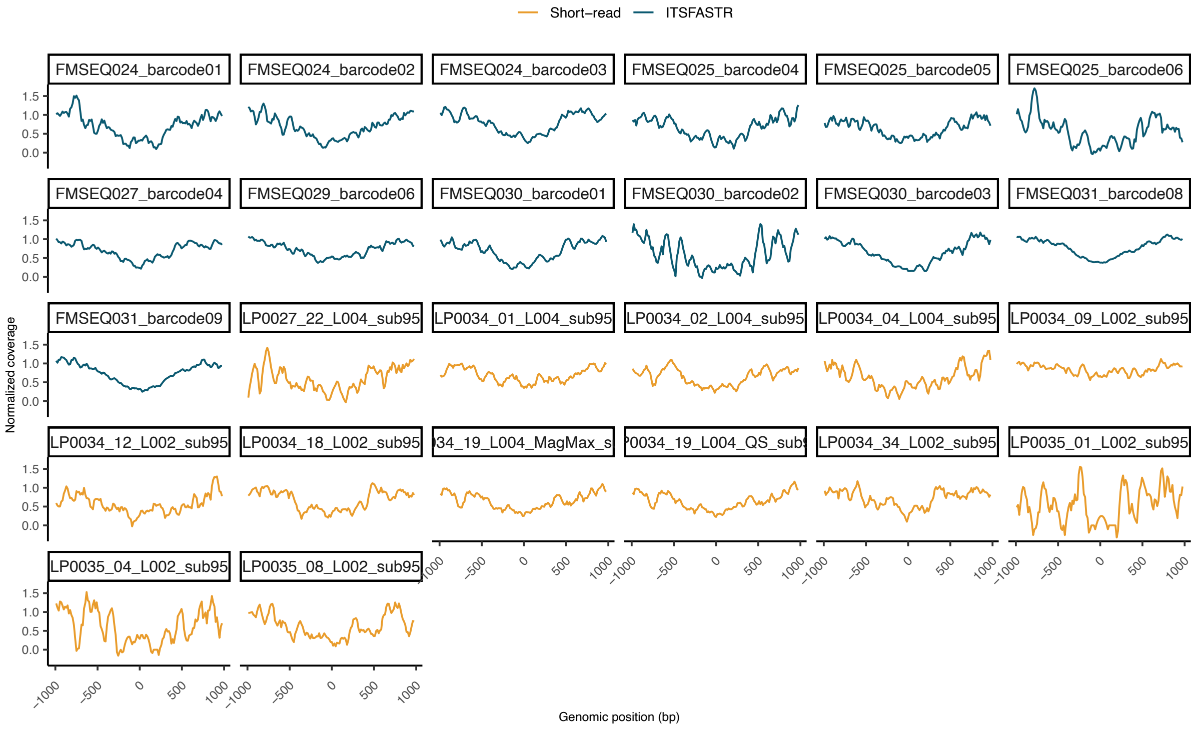


**Supplementary Figure 5: sequencing coverage near TSS regions of plasma samples.**

#### Supplementary Figure 6

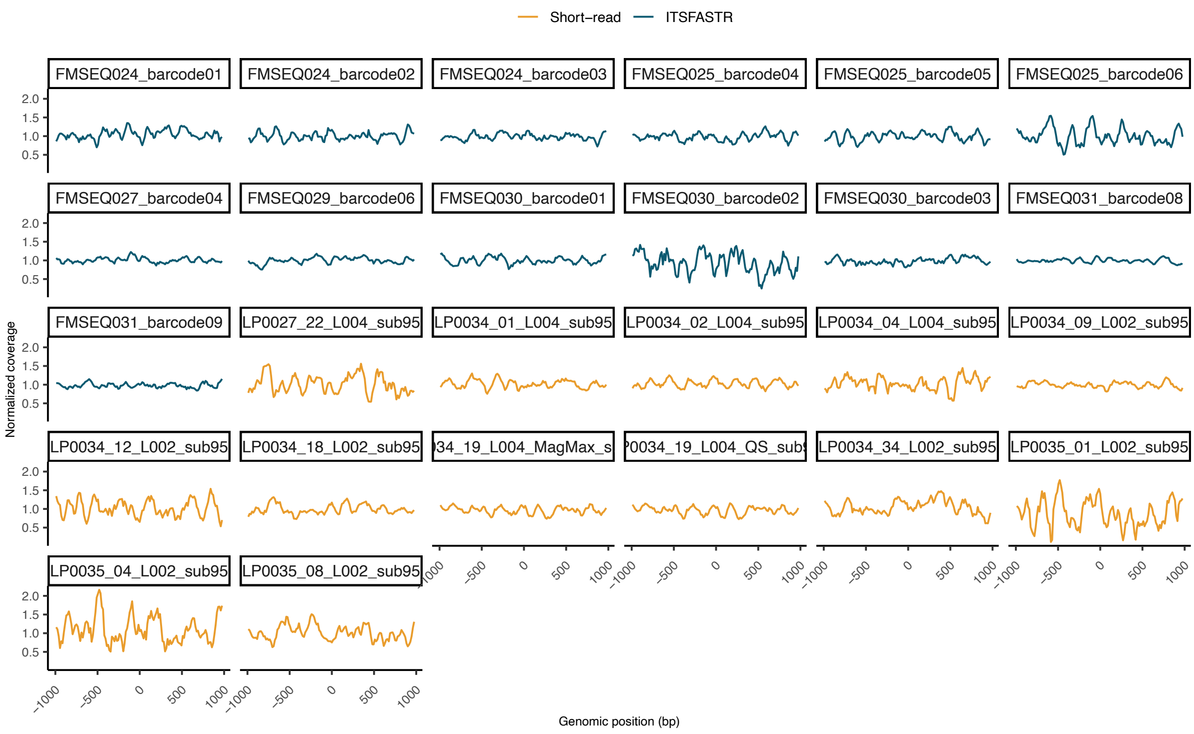


**Supplementary Figure 6: sequencing coverage near nucleosome-rich regions of plasma samples.**

#### Supplementary Figure 7

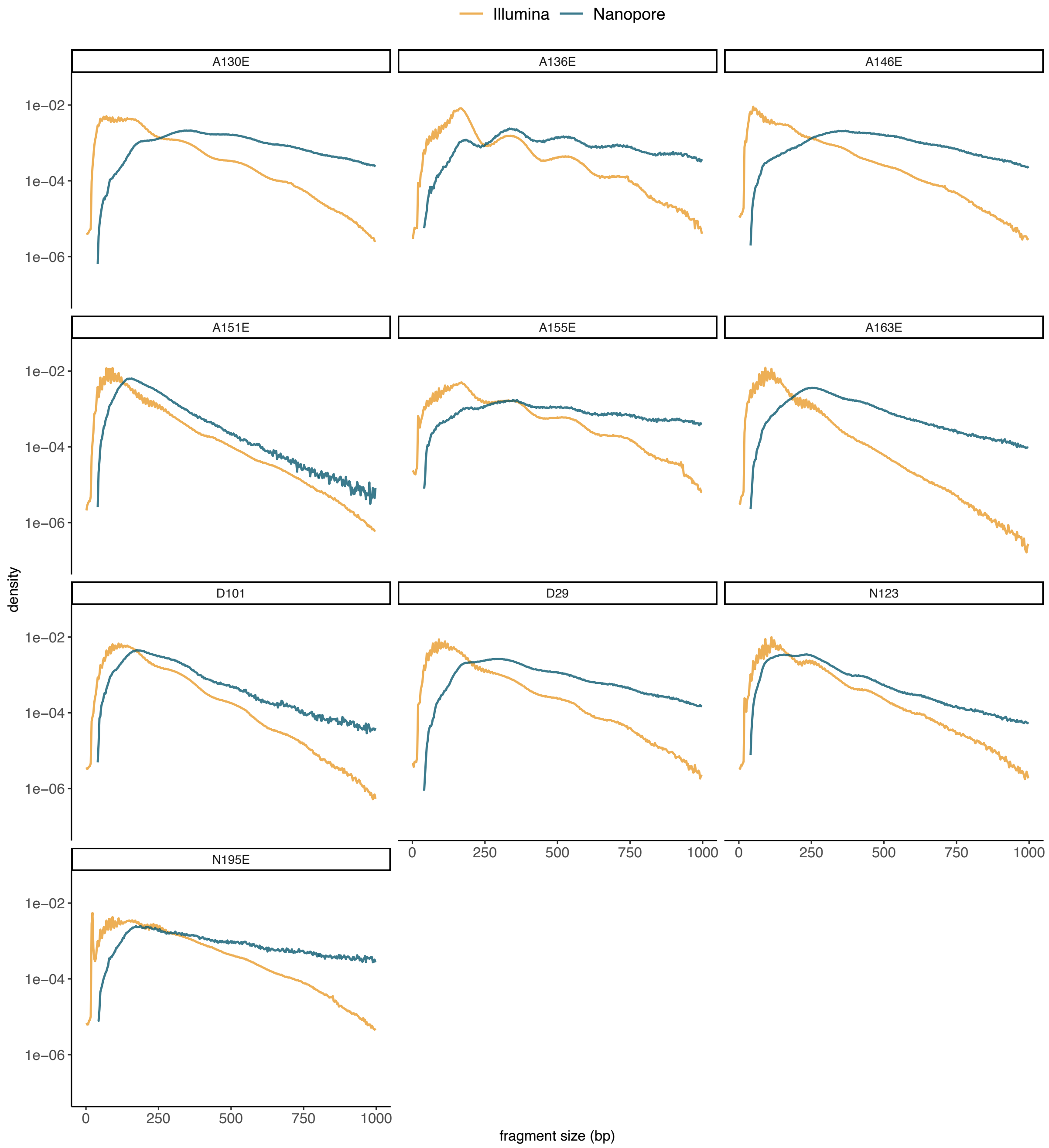


**Supplementary Figure 7. cfDNA fragment size distribution for the urine samples included in this study.** Color indicates the sequencing type (blue: Nanopore and orange: Illumina). Header numbers represent patient IDs.

#### Supplementary Figure 8

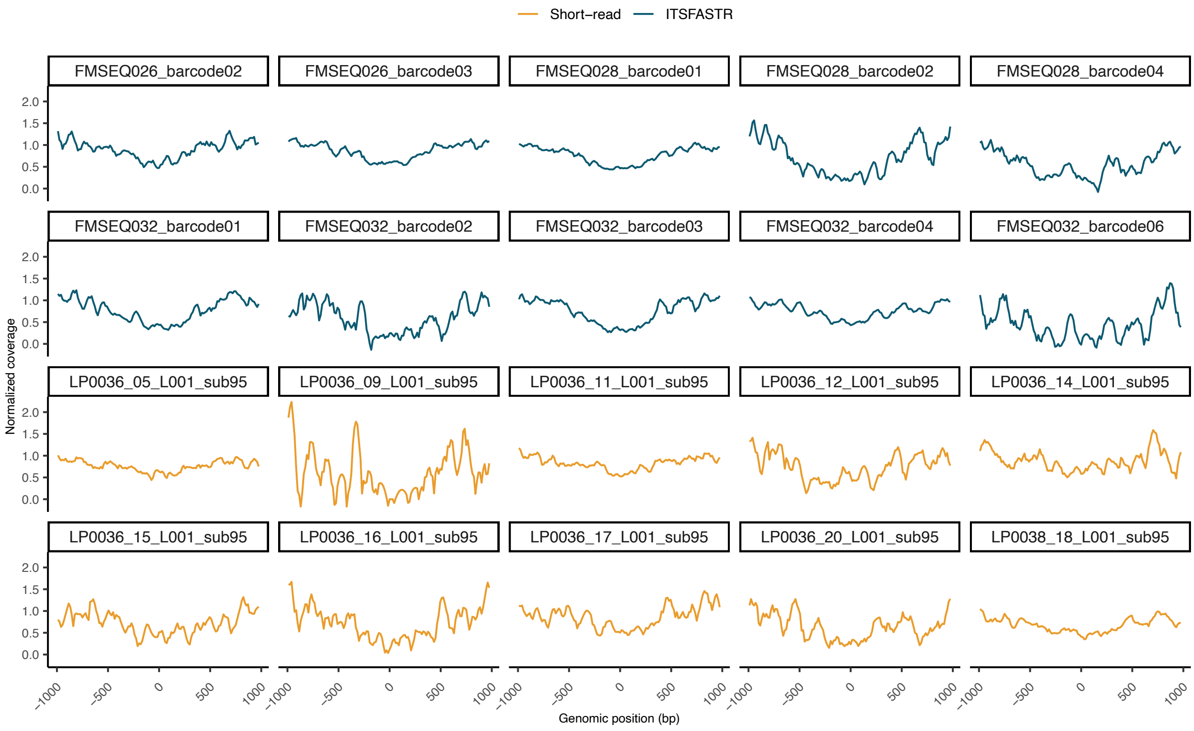


**Supplementary Figure 8: sequencing coverage near TSS regions of urine samples.**

#### Supplementary Figure 9

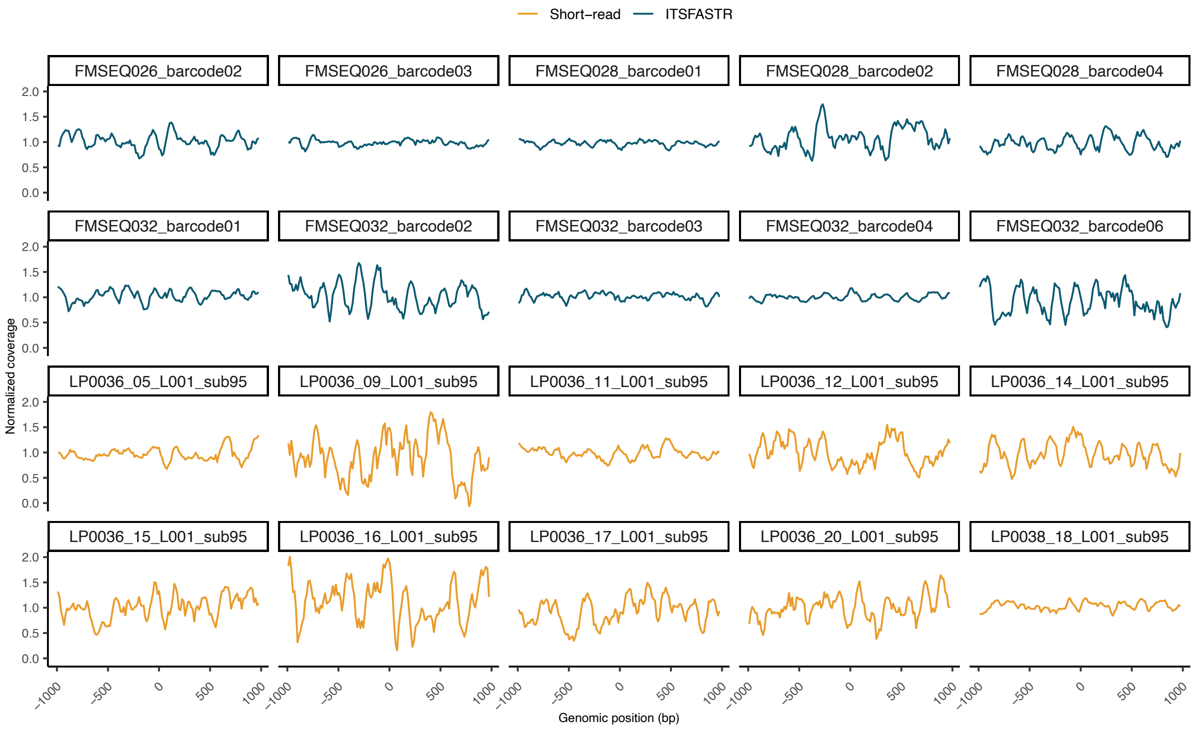


#### Supplementary Figure 6: sequencing coverage near nucleosome-rich regions of urine samples.
